# Demographics, Overlap, and Latency of Severe Cutaneous Adverse Reactions in an FDA Database

**DOI:** 10.1101/2025.03.05.25323441

**Authors:** Eric Milan Mukherjee, Dodie Park, Amir Asiaee, Matthew S. Krantz, Cosby A. Stone, Michelle Martin-Pozo, Elizabeth J. Phillips

## Abstract

**Importance:** Severe cutaneous adverse reactions (SCARs), including Stevens-Johnson syndrome/toxic epidermal necrolysis (SJS-TEN), drug reaction with eosinophilia and systemic symptoms (DRESS), acute generalized exanthematous pustulosis (AGEP), and generalized bullous fixed drug eruption (GBFDE), are rare but life-threatening drug hypersensitivity syndromes. Due to their low incidence and diagnostic complexity, large-scale characterization of SCAR is challenging.

**Objective:** To characterize the demographics, causative agents, trends, latency, and phenotypic overlap of SCAR using a large-scale, sanitized pharmacovigilance dataset from FAERS (FDA Adverse Event Reporting System).

**Design:** Cross-sectional study of spontaneous adverse event reports. Cases were drawn from the U.S. Food and Drug Administration Adverse Event Reporting System (FDA FAERS) from January 2004 to December 2023 and subjected to sanitization and deduplication. Disproportionality analysis was used to characterize causative agents. Machine learning (random forest classifiers) was used to analyze predictors of drug latency and mortality.

**Setting:** Global pharmacovigilance reports submitted to FAERS.

**Participants:** A total of 56,683 deduplicated SCAR reports were identified, representing 0.33% of reports during the study period.

**Exposures:** Suspected causative drugs, including both small molecules and biologics.

**Main Outcomes and Measures:** Main outcomes included the frequency and distribution of SCAR syndromes, reporting trends over time, latency from drug start to reaction onset, drug-specific disproportionality (PRR, ROR, IC), and co-reporting between SCAR types and related conditions.

**Results:** A total of 56,683 unique SCAR reports were identified, including SJS-TEN (28,871), DRESS (22,444), AGEP (6,183), and GBFDE (150). We identified 237 drugs with significant disproportionality for SCAR overall. Co-reporting between SCARs was significantly enriched (p < 10^-200^), suggesting overlapping phenotypes. Latency varied by drug and syndrome (median: GBFDE 3 days, AGEP 4 days, SJS-TEN 15 days, DRESS 24 days).

**Conclusions and Relevance:** SCAR syndromes display distinct but overlapping phenotypes, with variable latency and diverse causative agents. These findings, based on the largest SCAR dataset to date, highlight the need for improved classification frameworks and molecular validation. Large-scale pharmacovigilance, integrated with genomic and histopathologic data, will be critical to improving diagnosis, mechanistic understanding, and clinical management of SCAR.

## Introduction

Adverse drug reactions (ADR) are one of the top 10 causes of death in the developed world, costing up to $130 billion annually in the US.^1^ Among the most serious ADRs are severe cutaneous adverse reactions (SCARs), including Stevens-Johnson Syndrome/Toxic Epidermal Necrolysis (SJS-TEN), drug reaction with eosinophilia and systemic symptoms (DRESS), acute generalized exanthematous pustulosis (AGEP), and generalized bullous fixed drug eruption (GBFDE). These conditions are distinguished by morphology, causative agents, systemic involvement, and latency. For instance, AGEP and GBFDE manifests within days of exposure, while DRESS and SJS-TEN can take weeks.^2^

SCAR has incidence of 0.4 to 0.6 per million/year in the developed world, with mortality up to 70 percent in high-risk populations.^2^ SCAR can lead to signficant cutaneous damage, systemic involvement, and long-term sequelae. Because of their rarity, SCAR are difficult to study at scale. The largest published retrospective studies of SCAR have 300-400 cases, while the largest meta-analysis of SJS-TEN had 2917 patients.^3,4^ Given it’s rarity, there are many unanswered questions, including intermediate phenotypes between different SCAR, variations in onset and latency between causative agents, and geographic and temporal variation.

The FDA Adverse Event Reporting System (FAERS) is a valuable public database collecting information on ADRs. FAERS contains millions of reports submitted by providers, patients, and manufacturers. However, FAERS is hampered by unstructured drug names, duplication, and inconsistent detail.^5^ Understanding phenotype-specific latency distributions can guide discontinuation decisions in hospitalized patients. Likewise, knowledge of drug disproportionality profiles – both high-risk and unexpectedly low-risk agents – can refine differential diagnosis and influence prescribing practices in dermatology, oncology, infectious diseases, and primary care.

In this study, we sanitized FAERS and analyzed reports from 2004 to 2023, focusing on temporal trends, disproportionality, and overlap conditions. By pairing rigorous data cleaning with clinically oriented analyses, our goal was to generate findings that can be directly applied to patient care, supporting earlier recognition, more accurate classification, and more targeted management of these life-threatening reactions.

## Methods

Coding was done in RStudio 2024.12.0 Build 467 with R version 4.4.2, PyCharm 2024.2.0.1 with Python 3.12, PostgreSQL 16.3 (with PgAdmin IV), with GPT4o assistance. Details are available in the **Online Supplement**.

### Data

Quarterly ASCII files were fed into AEOLUS, which maps drugs and outcomes to standardized vocabularies and deduplicates by removing cases that match all four demographic fields (event date, age, sex and reporter country) using the previously published AEOLUS model.^6^ Unmapped drugs and outcomes were manually mapped by two researchers (EMM and DP), resulting in 100% mapping of rows in both tables.

### Disproportionality Analysis

To evaluate drug-event associations, we constructed 2×2 contingency tables for each drug–SCAR pair using unique patient identifiers. Patients exposed to a given drug (as primary suspect) and those with a reported SCAR were identified, and the intersection defined the count of cases (cell a). The remaining cells (b, c, and d) were derived based on the total number of unique patients across drug and event datasets. Disproportionality metrics – including proportional reporting ratio (PRR), reporting odds ratio (ROR), and Information Component (IC) based on Bayesian Confidence Propagation Neural Networks – were calculated with 95% confidence intervals, with formulas detailed in the online supplement. Chi-squared values with Yates’ correction were calculated using base R. We identified significant associations by evaluating whether the lower bound of the 95% confidence interval exceeded 1 for PRR and ROR, or 0 for IC (IC025).

### Here’s an expanded version you can drop into the Methods

#### Overlap Analysis

For overlap analyses, a patient was considered to have an overlap phenotype if their outcome data contained at least one preferred term from each of two different SCAR or cutaneous ADR phenotypes. For every pair of phenotypes, we counted the number of patients with each individual phenotype (N_A, N_B) and the number with both (N_overlap) and computed the expected overlap under independence as N_expected = (N_A×N_B) / N_total. From these quantities we derived: (i) lift (observed/expected overlap), (ii) the Jaccard index (N_overlap divided by the union of patients with either phenotype), and (iii) the phi coefficient as a measure of correlation between the two binary phenotypes. For overlaps between cutaneous ADRs, p-values were calculated using a one-sided binomial test (testing whether N_overlap exceeded the expectation under independence), with Benjamini–Hochberg correction for multiple testing. These metrics were summarized for SCAR–SCAR pairs and visualized for all cutaneous ADR pairs, including a ranked plot of the top overlaps by lift.

#### Time-To-Event (TTE) Analysis

Analyses were restricted to primary-suspect (PS) drugs. PS drug records were linked to the therapy table by (compositeid, drug_seq) to obtain start_dt; outcome records were linked with demographics to obtain event_dt. Rows were retained only when both dates were complete (YYYYMMDD), and TTE was computed as event_dt − start_dt (days). We applied phenotype-specific minimum TTE thresholds – SJS/TEN: exclude TTE ≤ 3 days; DRESS: exclude TTE ≤ 7 days; AGEP and GBFDE: exclude TTE ≤ 0 days – then windowed to 0 < TTE ≤ 180 days. To minimize repeated dosing artifacts, we kept the shortest TTE per patient×drug; for phenotype-level summaries we then kept the shortest TTE per patient. Values outside [Q1 – 1.5×IQR, Q3 + 1.5×IQR] were trimmed for plotting. Patients coded with multiple phenotypes (e.g., SJS/TEN and DRESS) appear in each relevant cohort.

## Results

### Demographics

After deduplication and data cleaning, a total of 56,683 SCAR cases were identified, representing 0.33% of all unique patient reports in FAERS from January 2004 to December 2023 (**eFigure 1**). This process began with over 20.8 million unique patient identifiers (PIDs) linked across the FAERS Reaction, Drug, Outcome, Demographics, and Therapeutics files. Drug and adverse event terms were mapped to standardized vocabularies using the AEOLUS method, followed by manual mapping of any unmapped entries (1.4% of drug rows and <0.01% of adverse event rows). Only fully mapped, deduplicated records were retained for downstream analyses, yielding 17.2 million unique PIDs with sanitized drug-outcome pairs.

Among these, the most frequently reported phenotype was SJS-TEN (28,871 cases), followed by DRESS (22,444), AGEP (6,183), and GBFDE (150). Temporal trends showed a steady increase in SCAR reporting over the study period, peaking in 2019. While SJS-TEN counts have remained relatively stable, DRESS reports have risen consistently, surpassing SJS-TEN as the most frequently reported SCAR annually since 2017.

Age statistically differs between SCAR, with GBFDE having lowest median (46 years, IQR [35-65]), then SJS-TEN (52 years, [30-68]), DRESS (52 years, [31-66]), and AGEP (59 years, [41-73]). Each SCAR shows female predominance except GBFDE. Analyzing sex and age (**eFigure 2**) shows that males have a bimodal age distribution, while female patients overtake males in adolescence before “catching up” in older age. Reports originate from 140 countries and territories (**eFigure 3 and 4**). USA (15607), France (9530), and Japan (5248) contributed the most reports. SJS-TEN outnumbers DRESS in all top 10 countries except for France, Spain, and Portugal, possibly due to trends in reporting and prescription. India reports the most GBFDE (44 cases), followed by France (28), the US (15) and Iran (15).

### Temporal Trends in Causative Agents

Using criteria from Evans et al (>2 cases, PRR ≥ 2, chi-squared ≥ 4), we find 237 drugs with positive signal when aggregating all SCAR together (**Supplemental Table**).^7^ When splitting by phenotype, there were 229 drugs with a positive signal for SJS-TEN, 161 for DRESS, 130 for AGEP, and 16 for GBFDE.

We analyzed temporal trends in SCAR reporting (**Figure 2**). The top five causative agents (allopurinol, vancomycin, lamotrigine, ibuprofen, and carbamazepine) are consistently responsible for ∼20% of new reports. The proportion of new reports due to biologics has steadily increased over time (p < 0.0001 by the Cochrane-Armitage test), comprising 8.2% of new reports in 2023, up from 2.6% in 2015 and 1.2% in 2004. Subdividing biologics by function, cases associated with immune checkpoint inhibitors (ICIs) have increased steadily since their 2011 approval (p < 0.0001, Cochrane-Armitage), comprising 49.1% of reports with a biologic as the primary suspect in 2023.

**Figure 1.**
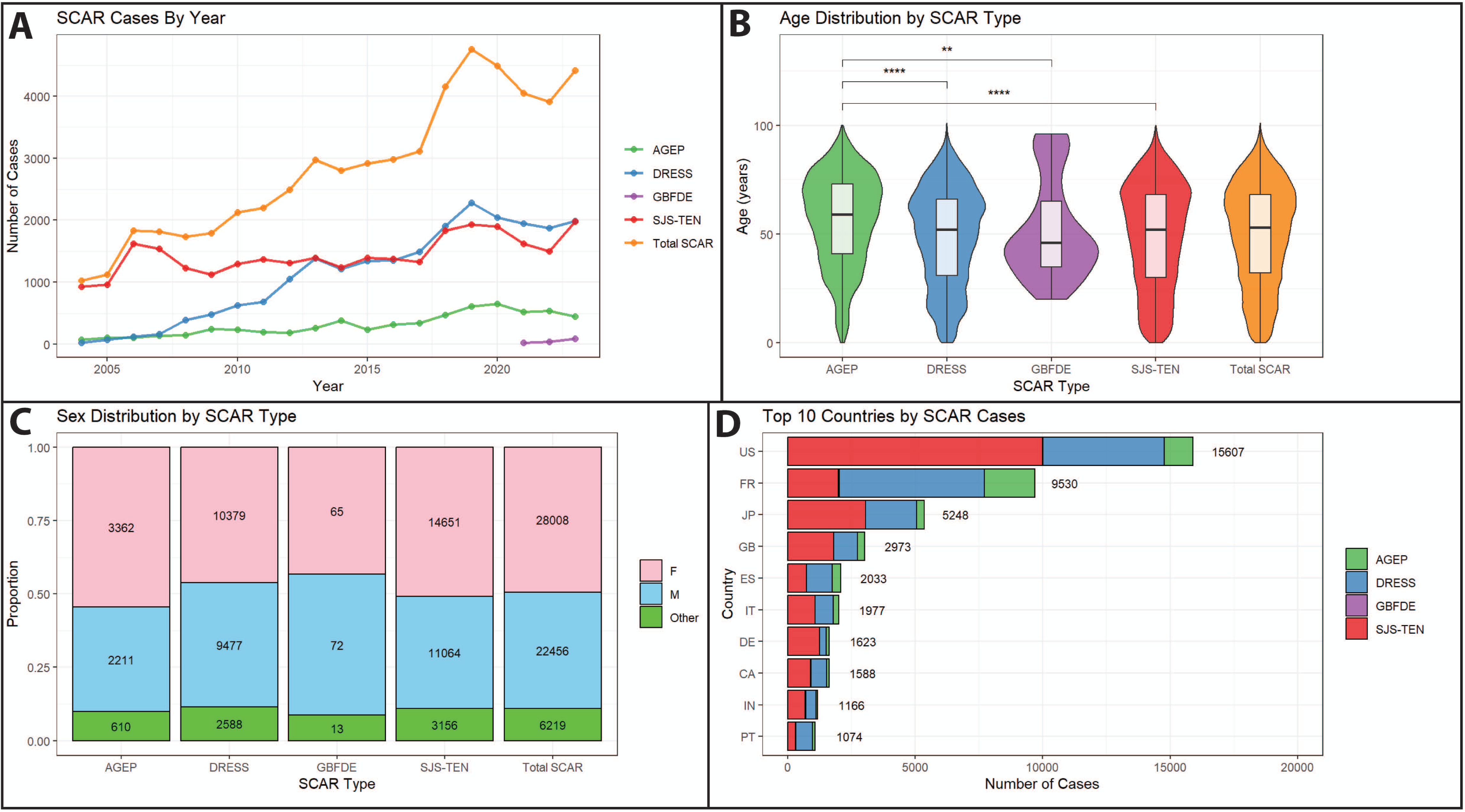
Demographic Summary of SCAR. (A) SCAR reporting over time in FAERS, by quarter. (B) Age distribution by SCAR. Pairwise comparisons done by Wilcoxon signed-rank test. **p < 0.01, ***p < 0.001, ****p < 0.0001. (C) Sex Distribution by SCAR. All patients without “M” or “F” sex markers were classified as “other”. (D) Top 10 Countries by SCAR cases. US - United States, FR - France, JP - Japan, GB - Great Britain, ES - Spain, IT - Italy, DE - Germany, CA - Canada, IN - India, PT - Portugal.

**Figure 2.**
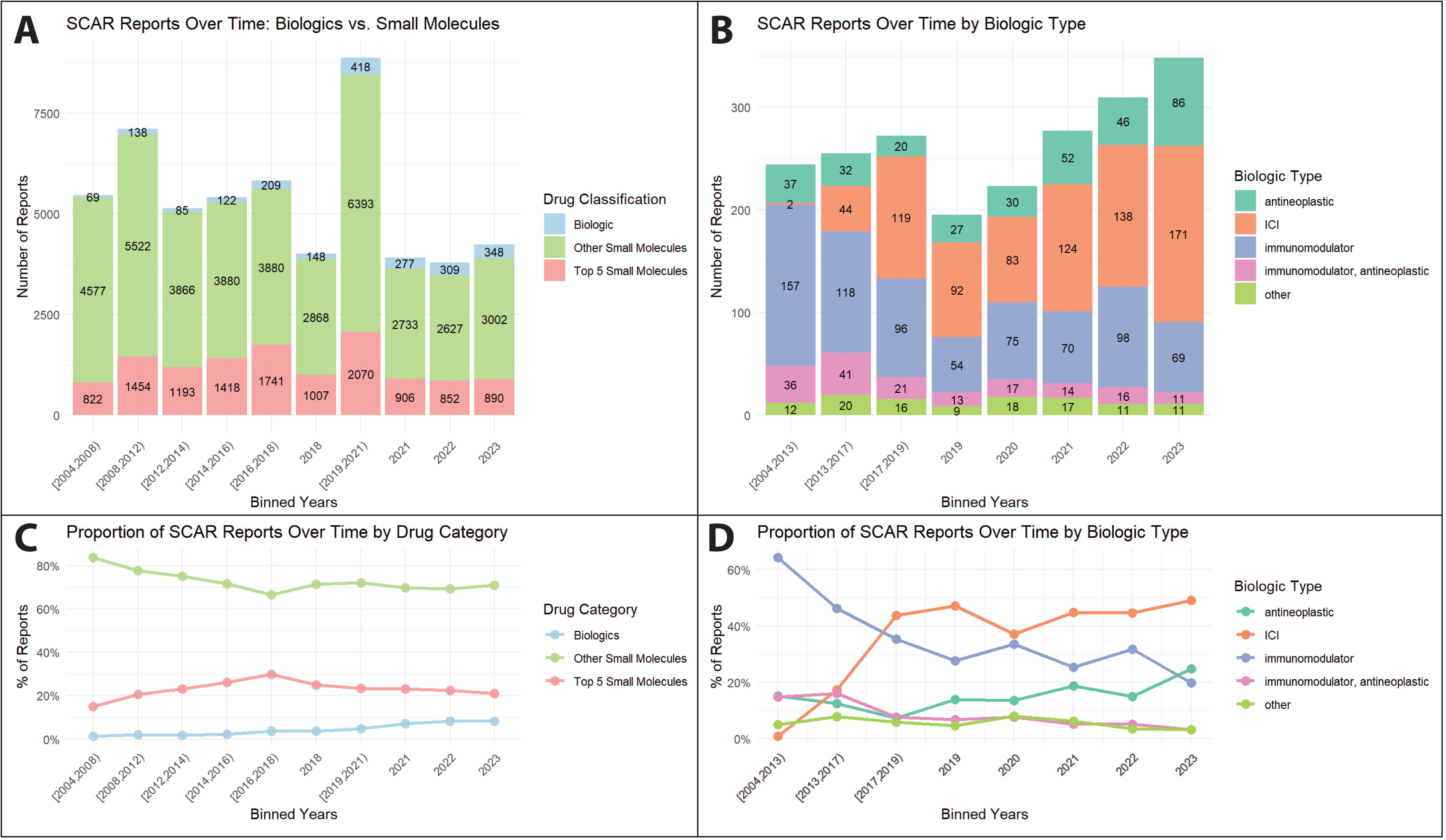
Trends in SCAR Reporting. Number of reports were aggregated by approximately-equal-frequency bins. (A) Number of SCAR reports whose primary cause is a “top 5” small molecule (allopurinol, carbamazepine, ibuprofen, lamotrigine, vancomycin), another small molecule, or a biologic. (B) Number of SCAR reports subdivided by type of biologic. ICI = immune checkpoint inhibitor. (C) Proportion of SCAR reports by drug category over time. Proportion of biologics has increased over time (p < 0.0001 by Cochrane-Armitage test). (D) Proportion of total biologic-related SCAR reports subdivided by type. Proportion of ICI has increased over time (p < 0.0001 by Cochrane-Armitage test).

### Disproportionality Analysis

**Figure 3** plots the information content (IC) of the top ≤20 drugs with >2 cases. **eFigure 4** shows the top 100 drugs when aggregating all SCAR together. Several well-known culprits (e.g. lamotrigine, allopurinol, and phenytoin for SJS-TEN, allopurinol, vancomycin, and lamotrigine for DRESS) are strongly associated (IC ≥ 3). Other, less-well-known associations (e.g., enfortumab and zonisamide for SJS-TEN, ciprofloxacin and valacyclovir for DRESS) have clear positive signals.^8–10^

**Figure 3.**
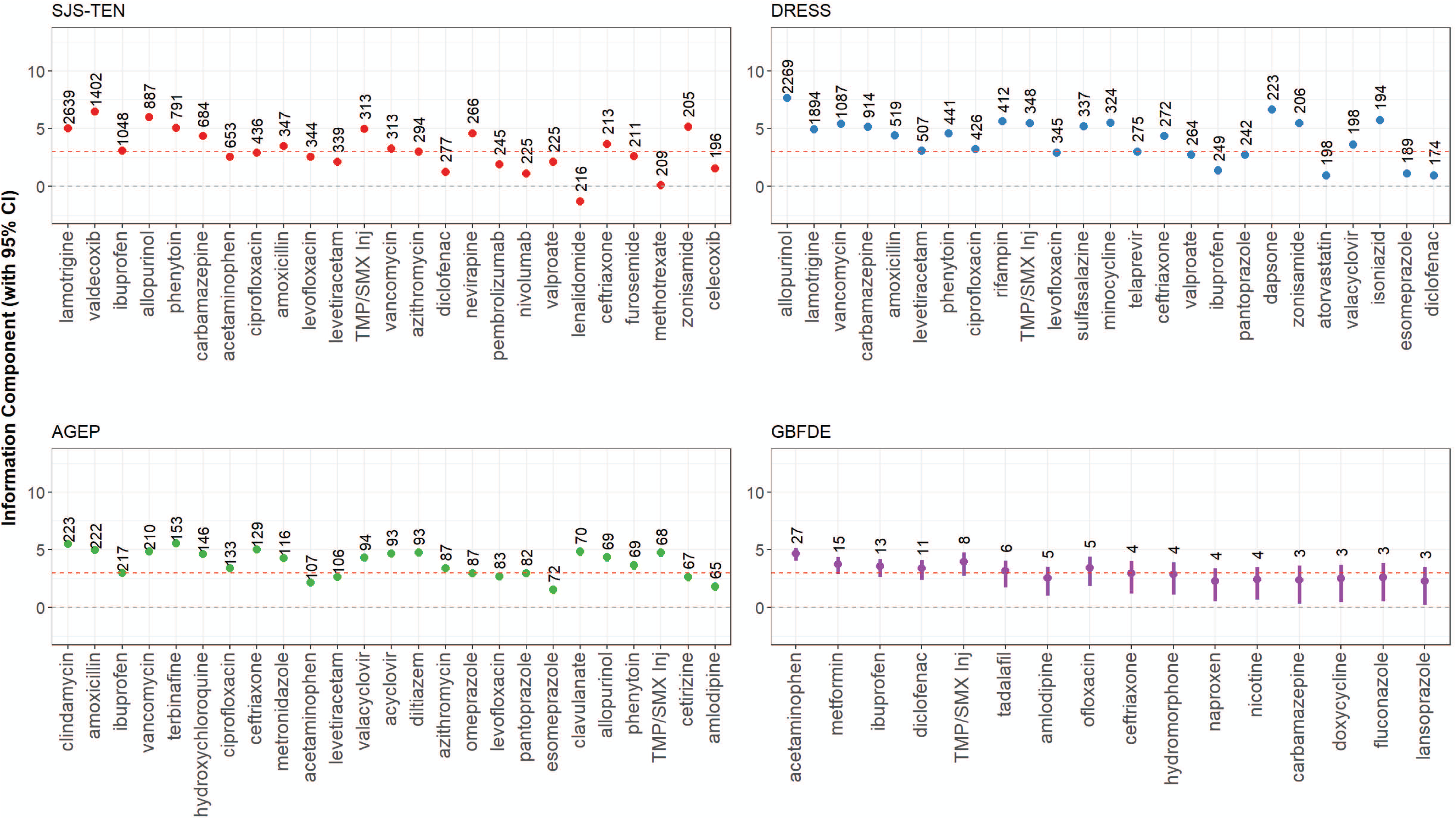
Disproportionality Analysis. Top <20 drugs for each SCAR by number of cases (with a minimum of 3 cases) are displayed. Each point represents the IC of a given drug-SCAR combination, with the total number of cases noted just above. Confidence intervals span IC025 to IC975. Red dashed line represents IC of 3, suggesting strong disproportionality.

### Disproportionate Co-Reporting

Overlap conditions between SCAR are rare, difficult to diagnose, and controversial in the literature.^14^ Disproportionality of co-occurring SCAR is shown in **Figure 4**. For SJS-TEN/DRESS, allopurinol (71 cases), lamotrigine (71), and phenytoin (49) are the most frequent suspects and have IC > 3. Co-reporting with AGEP is rarer; phenytoin (26 cases), ciprofloxacin (18), and allopurinol (14) are the most frequent culprits with co-reported DRESS, all with IC > 3, and phenytoin (34 cases), amlodipine (29), and ibuprofen (13) are the most frequent suspects of co-reported SJS-TEN.

**Figure 4.**
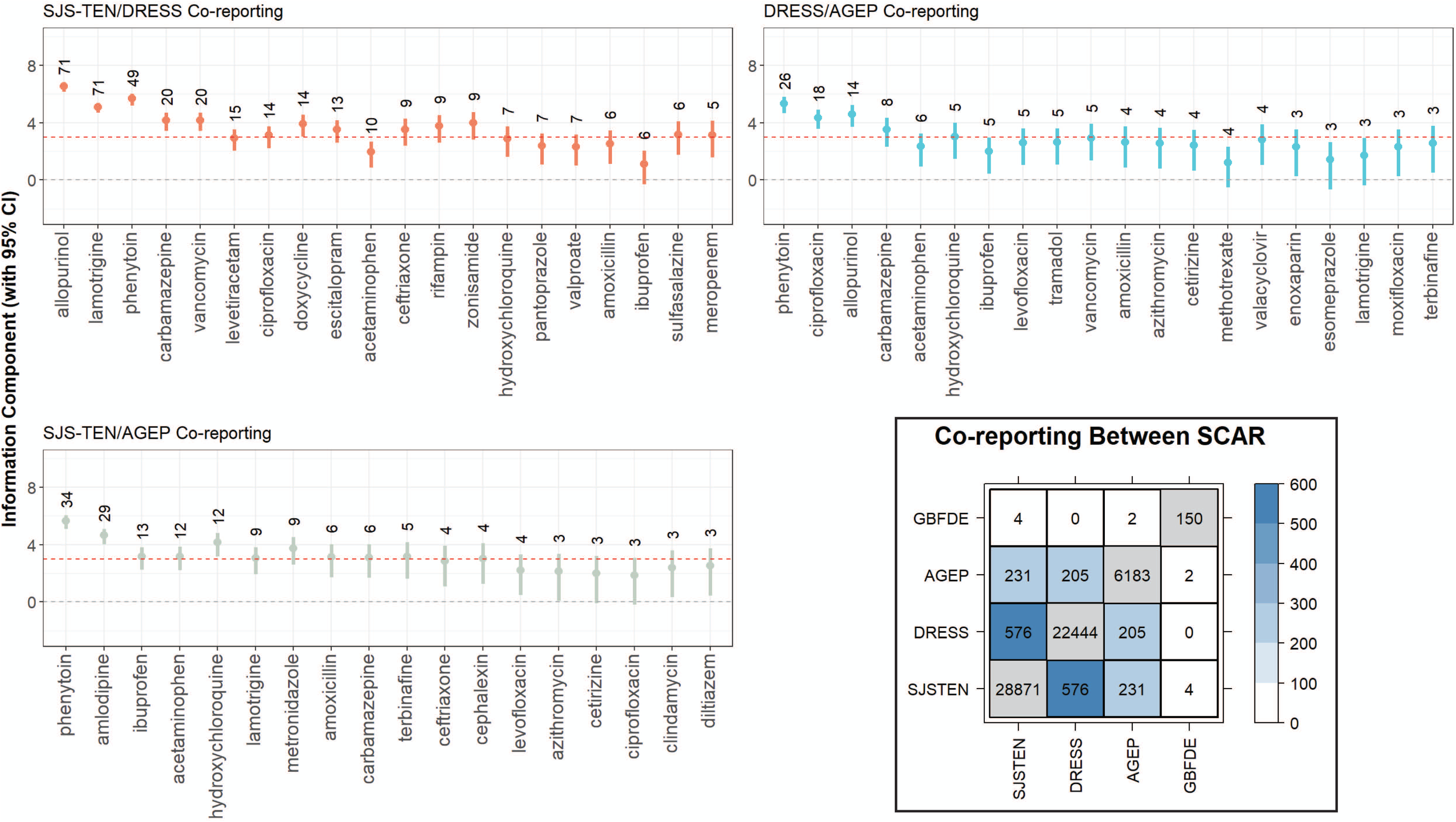
Disproportionality Analysis and Co-reporting of SCAR. Information component, with 95% confidence interval (IC025 to IC975) is plotted for all medications with at least 3 cases of co-reported SCAR. Inset: number of patients with co-reported SCAR, pairwise. Elements on the diagonal are the total patients with each SCAR.

Got you — here’s a corrected version of the two paragraphs with the **right N and p-values from the figure** and a short note about lift/Jaccard/phi folded in.

To better understand co-reporting patterns and to account for the possibility of misdiagnosis, we analyzed cases reporting more than one SCAR phenotype and examined co-reporting with other generalized cutaneous adverse drug reactions (ADR) that may mimic SCAR clinically. These phenotypes included autoimmune blistering disorders such as pemphigus and pemphigoid, symmetrical drug-related intertriginous and flexural exanthema (SDRIFE), fixed drug eruption (FDE), linear IgA bullous dermatosis (LABD), and systemic contact dermatitis (**eFigure 6**). Strong statistical enrichment was seen for overlap between SCAR phenotypes, including 576 cases of SJS–TEN with DRESS (p < 10^-300^; binomial test with Benjamini–Hochberg correction), 231 cases of SJS–TEN with AGEP (p = 2 × 10^-216^), and 205 cases of AGEP with DRESS (p = 1 × 10^-203^). For these SCAR–SCAR pairs, lift values were approximately 15–25 (observed/expected overlap), with Jaccard indices of only 0.7–1.1% and phi coefficients around 0.017–0.021, indicating that absolute overlap is rare but occurs far more often than expected under independence. These findings suggest that certain drug-induced eruptions either present with overlapping clinical and histopathologic features or are prone to diagnostic ambiguity in spontaneous reporting.

Co-reporting between SCAR and non-SCAR cutaneous ADR was less common but still notable in specific pairings. The strongest associations occurred between SJS–TEN and pemphigoid (63 cases, p = 5 × 10^-30^), SJS–TEN and LABD (23 cases, p = 3 × 10^-15^), and DRESS and LABD (43 cases, p = 2 × 10^-42^). Co-reporting was also enriched between pemphigus and pemphigoid (44 cases, p = 4 × 10^-36^) and between pemphigoid and LABD (14 cases, p = 4 × 10^-16^), consistent with known clinical and histologic overlap between these autoimmune blistering diseases. For these SCAR–non-SCAR and non-SCAR–non-SCAR pairs, lift values typically exceeded 10 and in some cases were >30, whereas Jaccard indices remained <1% and phi coefficients were in the ∼0.005–0.01 range. Thus, even when overlap is highly enriched statistically, co-reported phenotypes represent only a small fraction of all cases, highlighting the limitations of spontaneous reporting for conditions that require histopathologic confirmation and underscoring the need for harmonized clinical–pathologic definitions.

### Latency Varies by Causative Drug and SCAR

We conducted TTE analysis on a subset of cases filtered to those with full date information, filtered to those with time from starting the drug to event between 0 and 180 days, and subsequent filtering of outliers and removal of short latency cases (≤3 days for SJS/TEN, ≤7d for DRESS) to avoid protopathic bias (n = 11,584, **Figure 5A**). Latency differed by phenotype (p < 0.0001 by log-rank), with GBFDE having the shortest (3d, IQR [2-3], n = 5), followed by AGEP (4d, [2-9], n = 1541), SJS-TEN (15d, [9-26], n = 6111), and DRESS (24d, [15-35], n = 4047). Median latency varies substantially by causative drug (**Figure 5B**), though AGEP has faster onset for any given drug than SJS/TEN or DRESS to that same drug (**eFigure 7**). For SJS-TEN, NSAIDs (ibuprofen, acetaminophen) and antibiotics (ceftriaxone, amoxicillin, azithromycin) have short latency, while ICI (pembrolizumab, nivolumab), allopurinol, and anticonvulsants (zonisamide, phenytoin) have long latency. For DRESS, ibuprofen, amoxicillin, and the kinase inhibitor vemurafenib have short latencies, while the protease inhibitor telaprevir has a median latency of 48.5 days. For AGEP, a few drugs have long latency (vancomycin, hydroxychloroquine, terbinafine).

**Figure 5.**
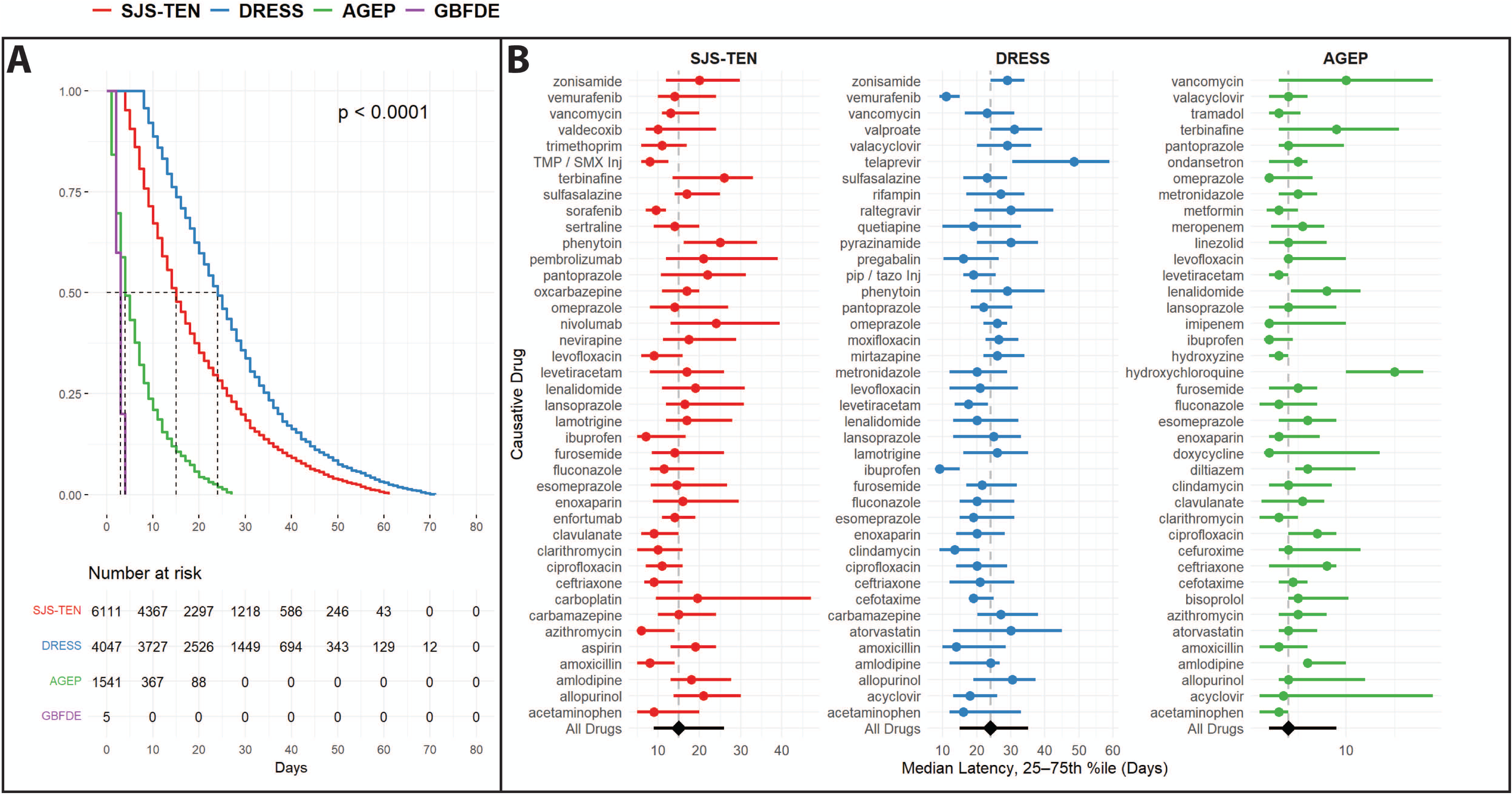
Drug Latency. (A) Survival curves show clear variation in median TTE/latency per SCAR. P-value by log-rank test. (B) Drug latency varies by causative drug, within each SCAR. The top 40 drugs (by number of cases) were plotted for each SCAR (GBFDE was excluded due to low number of cases). Each point represents median latency for that drug-SCAR combination, with IQR around the median. The “All Drugs” row for each SCAR shows the mean latency for each SCAR for all drugs (including those not displayed).

## Discussion

We report a comprehensive cross-sectional analysis of 56,683 SCAR cases from FAERS, 10-100x more than previous studies. SCAR reporting in FAERS has increased over time. For DRESS, this is likely due to the condition first being named in 1996, criteria being published in the mid-2000s, and FAERS first reporting in 2003.^15^ GBFDE, known since the 1980s, has only been reported in FAERS since 2021.^16^ While the plurality of reports are from high-resource nations, there are many reports from middle- and lower-income nations, reflecting SCAR’s global nature and the need for further global reporting.

Reported causative agents include well-known culprits like beta-lactams, anticonvulsants, and allopurinol, and less-studied agents like immunomodulators. The FDA’s Drug Safety-related Labeling Changes (SrLC) lists nine drugs since 2016 with label changes due to a link with SJS-TEN and/or DRESS – lamotrigine, allopurinol, phenytoin, carbamazepine, amoxicillin, trimethoprim-sulfamethoxazole (TMP-SMX), rifampin, levetiracetam, and vancomycin.^17^ Importantly, four of these drugs with the most label changes since 2016 for SCAR, DRESS, and/or SJS-TEN (allopurinol – 4 changes, phenytoin – 6 changes, rifampin – 5 changes, and TMP-SMX – 4 changes), are the same drugs that we found to be disproportionately represented in our data, providing evidence of the robustness of our methods but also indicating that these label changes may have impacted reporting.

The impact of these findings is underscored by the methods used to prepare FAERS, and by the breadth of medications studied. Prior studies have been limited by smaller sample sizes or focus on a few drugs.^18,19^ Additionally, prior studies have not been as thorough in sanitization, including limited deduplication efforts and lack of stringency in drug naming; this issue is well-documented and an area of active improvement.^5,20^

Even with our efforts, limitations remain. Because FAERS relies on voluntary reporting and lacks stringent diagnostic requirements (e.g., DRESS or AGEP cases do not require confirmation by RegiSCAR or EuroSCAR criteria), it cannot be used to estimate incidence, may lead to reporting biases, and remains susceptible to misdiagnosis. One source of error arises from confusion between phenotypically similar but etiologically distinct diseases, such as TEN-like pemphigoid, in which autoimmune blistering disorders mimic the widespread epidermal necrosis and mucosal involvement of SJS-TEN.^21^ Additional challenges include variable detail across reports – some omit concomitant drug data, exact dates, or full phenotyping – and the persistence of residual duplicate entries and typographical errors despite systematic cleaning. More advanced methods based on natural language processing and probabilistic grouping could further address these issues in future studies, alongside harmonized diagnostic criteria and the creation of specific MedDRA terms for emerging phenotypes.^22^

### Emerging Agents, Overlap Conditions and Mimickers

Particularly striking is the rise of ICI-induced SCAR, which presents a mechanistic challenge: whereas small molecules are thought to cause SCAR through direct perturbation of the T-cell receptor/peptide/MHC interface, ICIs may trigger reactions by releasing peripheral tolerance, resulting in a broad spectrum of immune-related adverse events, including SJS-TEN-like presentations.^23^ ICIs in our dataset were associated with longer latencies compared to traditional culprits, suggesting a distinct pathophysiology. It remains unclear whether these reactions represent true SJS-TEN or a distinct phenotype like PIRME.^24^ Additionally, ICIs may lower the threshold for small-molecule-induced SCAR, suggesting that some cases may be mis-attributed.^25,26^ A similar ambiguity is the antibody-drug conjugate enfortumab vedotin, which has been reported to cause both “classic” SJS-TEN and an SJS-TEN-like eruption histologically similar to toxic erythema of chemotherapy, likely due to direct toxicity on keratinocytes.^27^ Together, these findings suggest that immunotherapy-related mucocutaneous toxicity may span a spectrum that intersects, but is not identical to, classic SCAR.

Full phenotyping and histopathology are often required to diagnose conditions meeting the criteria of >1 SCAR. While such data is unavailable in FAERS, our work offers some insights. We demonstrate that co-reporting is higher than expected by random assortment and occurs more frequently than with other cutaneous conditions commonly misdiagnosed for each other (e.g. autoimmune bullous disease). This co-reporting may reflect either misdiagnosis, clinical ambiguity, or a true overlapping phenotype.

SJS-TEN/DRESS overlap, meaning cases meeting criteria for both, has been reported with allopurinol, anticonvulsants and TMP-SMX, among others.^28,29^ AGEP/DRESS overlap is more difficult. The condition is recognized as a discrete entity within FAERS, and in our data it appears in one case (primary suspect levofloxacin). Pustular DRESS is a known phenotype, with vancomycin, phenytoin, doxycycline and piperacillin-tazobactam implicated in a recent review (noting that these patients didn’t meet full AGEP criteria).^30^ A retrospective study of 216 patients found one probable case of DRESS-AGEP overlap meeting both criteria.^31^ Similarly, AGEP with TEN-like features, or AGEP/SJS-TEN overlap, is increasingly recognized.^32^ Many present with widespread desquamation and mucosal involvement appearing SJS-TEN-like, but with pathology showing pustules. Because histopathology is needed, many cases of both true AGEP/SJS-TEN overlap and AGEP with SJS-TEN-like features are likely misdiagnosed. Further harmonization of criteria and coordination are needed to characterize these cases.

### Causative Agents and Latency

Drug latency is critical in the management of SCAR, as different SCAR phenotypes differ significantly by latency, as reproduced in our data. In tandem with drug timelines, prior knowledge of latency is vital to determining the causative agent in complex cases. We have demonstrated that causative agents have a strong influence on latency, and that some agents have unusually long or short latencies, which is valuable knowledge in a clinical setting.

A few causative agents stand out in terms of unusually long or short latencies. The long latency of hydroxychloroquine- and terbinafine-induced AGEP has been described, in stark contrast to AGEP’s usual very short latency.^33,34^ SJS-TEN attributed to pembrolizumab and nivolumab also have longer latencies, consistent with the literature on PIRME.^24^ Vemurafenib has previously been attributed to rapid-onset DRESS.^35^ The reason for the dependence of latency on causative agent is not well-understood and bears further study, perhaps by considering chemical structure, metabolism, and time needed for antigen presentation.

A controversy in the field of SCAR is protopathic bias – namely, when a drug is misattributed as causative because it is prescribed early during SCAR to another drug. This is particularly notable for NSAIDs, which are often prescribed in the flu-like prodromal phase of SJS-TEN, and which have low latency in our data. An internal FDA review suggested acetaminophen caused rash, and mucosal erosion with a 1-3 day delay, though this finding is controversial.^36^ We attempted to filter out this effect by removing short-latency SJS/TEN and DRESS cases based on clinical experience; these cases may represent protopathic bias, accidental rechallenge, phenotypic misattribution (ex. Erythema multiforme vs SJS/TEN, or morbilliform drug eruption vs DRESS). Because GBFDE has only been an entity in FAERS since 2021, undoubtedly some SJS/TEN cases recorded before then were likely GBFDE. A concerted effort to confirm these cases with follow-up testing would be helpful.

### Conclusion and Future Directions

SCAR remains among the most devastating complications of pharmacotherapy, yet their rarity and complexity make them challenging to study in traditional clinical cohorts. Pharmacovigilance databases offer an opportunity to investigate SCAR at population scale, aggregating global data to identify trends, signals, and emerging culprits. In this study, we applied refined deduplication strategies and multi-dimensional analyses to characterize the epidemiology, latency, and phenotypic overlap of SCARs across more than 56,000 reports.

In parallel, the growing complexity of SCAR phenotypes demands new frameworks for classification that go beyond current syndrome labels. Emerging entities like PIRME, pustular DRESS, and AGEP with SJS-TEN-like features illustrate how existing diagnostic categories may fail to capture heterogeneity. Future efforts should prioritize harmonization of diagnostic criteria across SCAR, standardized integration of electronic health record information and histopathologic data, and adoption of probabilistic or spectrum-based models of classification. This will be essential not only for pharmacovigilance, but also for improving the design of trials, mechanistic studies, and treatment algorithms for these rare but life-threatening reactions.

## Data Sharing

Methods and data are available at https://github.com/capuhcheeno/SCARs_ICI-Manuscript-Scripts.

## Contributions

EMM conceived the project, wrote the manuscript, and conducted all analyses. DP conducted analysis and edited the manuscript. MMP, MSK, and CAS assisted with analysis and reviewed the manuscript. EP reviewed all analyses and the manuscript.

## Acknowledgements

E.J.P. is supported by the following grants from the National Institutes of Health (NIH): NIH U01AI154659, NIH P50GM115305, NIH R01HG010863, NIH R21AI139021, NIH R01AI152183, and NIH 2 D43 TW010559. E.J.P. is also supported by the National Health and Medical Research Council of Australia. E.M.M. is funded by a Vanderbilt University Medical Center internal career development award (Vanderbilt Faculty Research Scholars).

## Statement on Use of Artificial Intelligence

GPT4o was used to write, optimize, and debug R, SQL and Python code and improve language. Suggestions were cross-referenced in literature, package documentation, and resources like stackoverflow.com. All code and generated data were manually reviewed, tested, and verified by the authors. The authors take responsibility for all code generated and for all text.

## eFigure Legends

**eFigure 1. FAERS Deduplication and Sanitization.** Quarterly FAERS data files are deduplicated and combined, with drugs and outcomes mapped to standard terms. Any unmapped drugs and outcomes were manually mapped to standardized terms. From a data set of n = 17201564 unique patient IDs (PIDs), 56683 have at least one reported SCAR. Of those cases, 11,584 had sufficient data and were filtered for TTE analysis.

**eFigure 2. Age Versus Sex.** Age distribution, separated by sex, were determined for (A) total SCAR and (B) separated individual SCAR.

**eFigure 3. Geographic Distribution of SCAR.** Number of reports of all SCAR are displayed.

**eFigure 4. Geographic Distribution of Individual SCAR.** Number of reports of individual SCAR are displayed

**eFigure 5. Disproportionality Analysis of SCAR, Aggregated.** Information component, along with 95% confidence interval (from IC025 to IC975) were calculated for the top 100 drugs by number of cases. Red dashed line at IC = 3 indicates strongly disproportionate associations.

**eFigure 6. Disproportionate Co-reporting Between Cutaneous Adverse Reactions.** (A) Heatmap of pairwise co-reporting between cutaneous ADRs. The diagonal shows the total number of patients reported with each phenotype. Upper-triangle cells show, for each pair, the number of patients who had both reactions (N) and the p-value from a one-sided binomial test with Benjamini–Hochberg correction. (B) Overlap effect sizes for all 45 phenotype pairs, plotted as Jaccard index (fraction of the union of patients with both events) versus the phi coefficient (correlation between the two binary phenotypes). A small number of pairs – most notably SJS-TEN with DRESS and AGEP, and GBFDE with FDE – show modest but statistically robust positive association. (C) Horizontal bar plot of the top 20 cutaneous ADR pairs ranked by lift (observed / expected overlap under independence). Most pairs have lift values well above 1, indicating substantial enrichment of co-reporting, but these overlaps remain numerically rare, consistent with largely distinct patient populations for each phenotype.

**eFigure 7. Median time-to-event (days) with interquartile range (IQR) for selected culprit drugs across SCAR phenotypes.** Colored points mark medians and horizontal bars the IQR for SJS/TEN (red), DRESS (blue), and AGEP (green). Across many agents, DRESS shows longer median latency, with AGEP generally shortest.

