## Supplementary material for "Demographics, Overlap, and Latency of Severe Cutaneous Adverse Reactions in an FDA Database": Online Methods

ONLINE SUPPLEMENTAL METHODS

**DATA HANDLING**

*Processing of Raw Data*

The data processing pipeline was developed on a Linux system using Cygwin 3.5.3 on Windows. Python scripts were executed through the PyCharm client, while PostgreSQL 16.3, managed via PgAdmin IV, served as the database platform. OHDSI Usagi was used for mapping unmapped drugs, and Notepad++ was used for text editing. All scripts and supplementary files are hosted in the SCARs_ICI-Manuscript-Scripts GitHub repository: <https://github.com/capuhcheeno/SCARs_ICI-Manuscript-Scripts>

*Reference Data Preparation*

The schema faers was created to store FDA Orange Book and other reference data. The FDA Orange Book was downloaded from its official website and loaded using load_nda_table.sql. Additional reference tables, including country codes and EU drug names with active ingredients, were added to the schema using respective SQL scripts.

A second schema, cdmv5, was created to house OHDSI CDMV5 vocabulary tables. These vocabularies were downloaded from OHDSI Athena, including the MedDRA MSSO vocabulary, and loaded into PostgreSQL using Python scripts. Mapping tables, such as those linking MedDRA and SNOMED, were generated using SQL to facilitate downstream analyses.

*FAERS Data Preparation*

Current and legacy FAERS datasets were downloaded from the FDA website. For current data, files were processed and combined using shell scripts that added filename columns to each dataset (e.g., demographics, drugs, outcomes). These consolidated files were loaded into PostgreSQL using SQL scripts corresponding to each dataset type. For legacy data, additional manual steps were required to include datasets from 2004 Q4 and 2005 Q3, which were not available in the automated download. Legacy data files were similarly processed with scripts to consolidate and load them into PostgreSQL. Data inconsistencies, such as formatting issues in legacy demographics files, were corrected using Notepad++ before loading.

*Processing Steps*

To ensure unique case entries, duplicate records were removed using derive_unique_all_case.sql. Drug names were mapped to RxNorm using OHDSI Usagi. Unmapped codes in standard_case_drug.csv were exported from PostgreSQL, edited for compatibility, and manually mapped in Usagi. The mapped codes were re-imported and integrated into the database using scripts that standardized the drug mappings to create standard_case_drug_with_names.csv.

**DATA SANITIZATION**

*Drug and Outcome Tables*

Data from AEOLUS output, in particular the standard_case_drug_with_names.csv and standard_case_outcome.csv were manually cleaned as described below and transformed as follows (with snippets of the table for illustration):

standard_case_drug_with_names (AEOLUS output):

|  | **primaryid** | **Isr** | **drug_seq** | **role_cod** | **drug_concept_id** | **drug_name** |
| --- | --- | --- | --- | --- | --- | --- |
| 1 | 100033001 | *NA* | 1 | PS | 1512480 | Ibandronate |
| 2 | 100033011 | *NA* | 1 | PS | 1177480 | Ibuprofen |
| 3 | 100033021 | *NA* | 1 | PS | 734354 | Pregabalin |
| 4 | 100033042 | *NA* | 1 | PS | 1738521 | Doxycycline |
| 5 | 100033042 | *NA* | 2 | SS | 1103314 | Tramadol |
| … | | | | | |  |
| 54977140 | *NA* | 7910914 | 1018033938 | C | 1557272 | Alendronate |
| 54977141 | *NA* | 7910914 | 1018033940 | C | 974166 | Hydrochlorothiazide |
| 54977142 | *NA* | 7910915 | 1018033941 | PS | 735843 | Natalizumab |
| 54977143 | *NA* | 7910916 | 1018033942 | PS | 735843 | Natalizumab |

Is transformed to sanitized_drug_data:

|  | **compositeid** | **drug_seq** | **role_cod** | **drug_concept_id** | **drug_name** |
| --- | --- | --- | --- | --- | --- |
| 1 | pid.100033001 | 1 | PS | 1512480 | ibandronate |
| 2 | pid.100033011 | 1 | PS | 1177480 | ibuprofen |
| 3 | pid.100033021 | 1 | PS | 734354 | pregabalin |
| 4 | pid.100033042 | 1 | PS | 1738521 | doxycycline |
| 5 | pid.100033042 | 2 | SS | 1103314 | tramadol |
| … | | | | | |
| 54977140 | isr.7910914 | 1018033938 | C | 1557272 | alendronate |
| 54977141 | isr.7910914 | 1018033940 | C | 974166 | hydrochlorothiazide |
| 54977142 | isr.7910915 | 1018033941 | PS | 735843 | natalizumab |
| 54977143 | isr.7910916 | 1018033942 | PS | 735843 | natalizumab |

standard_case_outcome (AEOLUS output):

|  | **primaryid** | **Isr** | **Pt** | **outcome_concept_id** | **snomed_outcome_concept_id** |
| --- | --- | --- | --- | --- | --- |
| 1 | 100033001 | *NA* | Arthralgia | 3656812 | *NA* |
| 2 | 100033001 | *NA* | Diarrhoea | 35708093 | *NA* |
| … | | | | | |
| 45332774 | NA | 590503 | PALPITATIONS | 35205034 | *NA* |
| 45332775 | NA | 590503 | POST PROCEDURAL COMPLICATION | 36211677 | *NA* |

Is transformed to sanitized_outcome_data:

|  | **compositeid** | **pt** | **outcome_concept_id** |
| --- | --- | --- | --- |
| 1 | pid.100033001 | Arthralgia | 3656812 |
| 2 | pid.100033001 | Diarrhoea | 35708093 |
| … | | | |
| 45332774 | isr.590503 | PALPITATIONS | 35205034 |
| 45332775 | isr.590503 | POST PROCEDURAL COMPLICATION | 36211677 |

Unmapped outcomes (40 total) and unmapped drugs (1780 total) were exported for further inspection. Mapping failures were due to alternate names and typographical errors, which we manually corrected using the RxNorm and MedDRA databases. This resulted in 100% mapping outcomes. After mapping was complete, a few other modifications were made to shorten names (e.g. changing “hydrochlorothiazide” to “HCTZ”) and reduce redundancy – in particular, "piperacillin / tazobactam Injection" and "piperacillin / tazobactam Injectable Solution" were mapped separately, and were combined into one ID.

After all sanitization, sanitized_drug_data had 56389500 rows, representing 16551079 patients and 5492 drugs. sanitized_outcome_data had 51135682 rows, representing 17201516 patients and 23176 outcomes. Between these two tables, there are 17201564 unique patients, which is the total number of patients used for all calculations. Both tables were saved as .csv files for subsequent analysis.

*Demographic Information*

All DEMO files were combined into a single table, filtering to the latest case version. Age was harmonized to years (rather than weeks, days, hours, minutes) mathematically based on a 365-day and 52.1775-week year. Ages less than 0 years and over 120 years were replaced with NA values. Country of origin was filtered down to the country in which the reaction occurred (occr_country) or, if not available, the home country of the reporter (reporter_country). Full-length country names in legacy LAERS data were manually mapped to ISO 3166-1 alpha-2 two-letter country codes.

Further deduplication of rows with identical compositeids was conducted as follows (in order):

1. Replacing NA values in one duplicate with non-NA values in the other for any column
2. Keeping later event dates (event_dt), FDA dates (fda_dt) and reporting dates (rept_dt) when multiple dates exist
3. Keeping earlier filenames (which report which quarterly report the entry is taken from)
4. Replacing “NS” (not specified) sex values with “M” or “F” when available in another duplicate row.
5. Randomly selecting one of two conflicting rows (required for < 50 rows of 20750363 total)

This was subsequently saved as sanitized_demographics.csv.

*Drug Therapy Start and End Dates*

Drug administration times are stored in the THER tables from FAERS. These were combined and harmonized into a single sanitized_ther.csv table.

*Temporal Trends*

Temporal trends (Figure 3) were calculated by considering the filename of origin of a given case (which encodes the year and quarter). A manually-curated dictionary of biologics (biologic_drug_list.csv) was used to separate biologic medications from small molecules in this analysis. The top 5 drugs were defined as vancomycin, allopurinol, lamotrigine, carbamazepine, and ibuprofen, all common culprits of SCAR. Trends were analyzed using logistic regression and the Cochrane-Armitage test.

**CONTINGENCY TABLES AND DISPROPORTIONALITY ANALYSIS**

This analysis can be found in the R markdown file FAERS_SCAR_Analysis.Rmd

*Selection of Cutaneous Adverse Events from MedDRA terms*

The following mapping between our outcomes of interest and the MedDRA terms in the FAERS database was used:

| **SCAR or Condition** | **MedDRA Terms** |
| --- | --- |
| SJS-TEN | “Stevens-Johnson syndrome”, “SJS-TEN overlap”, “Toxic epidermal necrolysis” |
| DRESS | “Drug reaction with eosinophilia and systemic symptoms”, “Drug rash with eosinophilia and systemic symptoms”, “AGEP-DRESS overlap” |
| AGEP | “Acute generalised exanthematous pustulosis”, “AGEP-DRESS overlap” |
| GBFDE | “Generalised bullous fixed drug eruption” |
| Pemphigoid | “Pemphigoid”, “Ocular pemphigoid”, “Mucous membrane pemphigoid”, “Lichen planus pemphigoides”, “Pemphigoid antibody panel” |
| Pemphigus | “Pemphigus”, “Paraneoplastic pemphigus”, “Pemphigus disease area index” |
| SDRIFE | “Symmetrical drug-related intertriginous and flexural exanthema” |
| Systemic Contact Dermatitis | “Systemic contact dermatitis” |
| Linear IgA Disease | “Linear IgA disease” |
| FDE | “Fixed drug eruption”, “Fixed eruption” |

Abbreviations: Stevens-Johnson Syndrome (SJS), Toxic Epidermal Necrolysis (TEN), Drug Reaction with Eosinophilia and Systemic Symptoms (DRESS), Acute Generalized Exanthematous Pustulosis (AGEP), Generalized Bullous Fixed Drug Eruption (GBFDE), Symmetrical drug-related intertriginous and flexural exanthema (SDRIFE), Fixed Drug Eruption (FDE).

SJS-TEN, DRESS, AGEP, and GBFDE were grouped into the larger category of “SCAR” for some analyses.

*Contingency Tables*

Each drug-event pair was structured as a 2x2 contingency table:

|  | ADE Present | ADE Absent |
| --- | --- | --- |
| Drug Present | a | b |
| Drug Absent | c | d |

Thus:

- **a**: Number of cases reporting both the drug and the event.
- **b**: Number of cases reporting the drug but not the event.
- **c**: Number of cases reporting the event but not the drug.
- **d**: Number of cases reporting neither the drug nor the event.

We found that the original AEOLUS method produces contingency tables with an inconsistent total number of patients, suggesting double- or-undercounting. Thus, contingency tables for each drug-outcome pair were constructed as follows. First, all unique patient IDs exposed to a particular drug (restricted by role as appropriate) were obtained, and all unique patient IDs with a particular outcome of interest were obtained. The intersection of these two lists gave the number of cases (“a” in the contingency table). The other entries in each contingency table were computed by subtracting from the total number of patients exposed to the drug (as primary suspect), the total number of patients with the given outcome, and the total number of patients as appropriate.

| **Variable** | **Narrative Explanation** | **Calculation** |
| --- | --- | --- |
| Exp | Set of unique patients exposed to drug | Obtained directly from drug_data |
| Out | Set of unique patients with outcome | Obtained directly from outcome_data |
| Tot | # unique patients total | # unique patients in {drug_data ∪ outcome_data} |
| a | # unique patients exposed to drug WITH outcome | \|Exp ∩ Out\| |
| b | # unique patients exposed to drug WITHOUT outcome | \|Exp\| – a |
| c | # unique patients NOT exposed to drug WITH outcome | \|Out\| – a |
| d | # unique patients NOT exposed to drug WITHOUT outcome | Tot – (a+b+c) |

We utilized disproportionality analysis methods to evaluate drug-event associations, including Proportional Reporting Ratios (PRR), Reporting Odds Ratios (ROR), and Bayesian Information Components (IC) with 95% confidence intervals. A contingency table-based approach was applied to calculate these metrics for each drug-event pair.

*Proportional Reporting Ratio (PRR)*

PRR was calculated as:

PRR=a/(a+b)c/(c+d)PRR = \frac{a / (a + b)}{c / (c + d)}

95% Confidence Intervals (CIs) for PRR were calculated as:

PRR_{lower} = e^{ln(PRR) - 1.96 \sqrt{\frac{b}{a(a+b)} + \frac{d}{c(c+d)}}}

PRR_{upper} = e^{ln(PRR) + 1.96 \sqrt{\frac{b}{a(a+b)} + \frac{d}{c(c+d)}}}

*Reporting Odds Ratio (ROR)*

ROR was computed as:

ROR = \frac{ad}{bc}

95% Confidence Intervals (CIs) for ROR were calculated as:

ROR_{lower} = e^{ln(ROR) - 1.96 \sqrt{\frac{1}{a} + \frac{1}{b} + \frac{1}{c} + \frac{1}{d}}}

ROR_{upper} = e^{ln(ROR) + 1.96 \sqrt{\frac{1}{a} + \frac{1}{b} + \frac{1}{c} + \frac{1}{d}}}

*Information Component (IC)*

Bayesian shrinkage estimates for disproportionality were computed using the Information Component (IC) and 95% confidence intervals (IC025 and IC975).

The expected value (E) was calculated as:

E = \frac{(a + b)(a + c)}{a + b + c + d}

IC was calculated as:

IC = log_2 \left(\frac{a + 0.5}{E + 0.5}\right)

The lower (IC025) and upper (IC975) confidence intervals were calculated using simplified formulas as previously described[1]:

IC025 = log_2 \left(\frac{a + 0.5}{E + 0.5}\right) - 3.3 \cdot (a + 0.5)^{-1/2} – 2 \cdot (a + 0.5)^{-3/2}

IC975 = log_2 \left(\frac{a + 0.5}{E + 0.5}\right) + 2.4 \cdot (a + 0.5)^{-1/2} – 0.5 \cdot (a + 0.5)^{-3/2}

*Chi-Squared Test with Yates' Correction*

All calculations were implemented in R using built-in functions and the chisq.test() method with Yates' correction:

χ2=(∣ad−bc∣−0.5)2⋅(a+b+c+d)(a+b)(c+d)(a+c)(b+d)\chi^2 = \frac{(|ad - bc| - 0.5)^2 \cdot (a + b + c + d)}{(a + b)(c + d)(a + c)(b + d)}

*Overlap Analysis*

For overlap analyses (Figure 4), a patient was considered overlapping if their outcome data had at least one term from each of two different SCAR. For overlaps between cutaneous ADR (Figure S5), p-values were calculated using the binomial test (assuming a random draw from the entire FAERS database), with Benjamini-Hochberg correction for multiple testing.

**LATENCY AND TIME-TO-EVENT ANALYSIS**

*Drug Latency Analysis*

This analysis is found in the markdown file FAERS_TTE_Revamp.Rmd

We restricted analyses to primary-suspect (PS) drugs and used base R merge() to link PS drug records to the therapy table by (compositeid, drug_seq) to obtain start_dt, and to link outcomes with demographics to obtain event_dt (deduplicated to avoid .x/.y suffixes). Rows were retained only when both dates were complete in YYYYMMDD format, and time-to-event (TTE) was computed as event_dt - start_dt (days). Of 17,201,564 unique compositeid in the sanitized dataset, 4,070,635 (23.7%) had sufficient information to calculate TTE. We then created phenotype-specific subsets and wrote them to disk: final_table_TTE_SJSTEN.csv, final_table_TTE_DRESS.csv, final_table_TTE_AGEP.csv, final_table_TTE_GBFDE.csv, and an aggregate final_table_TTE_SCAR.csv containing all rows with any SCAR phenotype. Patients coded with multiple phenotypes (e.g., SJS/TEN and DRESS) appear in each relevant subset.

Filtering was used in all figures/analyses. After assembling TTE, we applied phenotype-specific minimum –

SJS/TEN: exclude TTE ≤ 3 days; DRESS: exclude TTE ≤ 7 days; AGEP and GBFDE: exclude TTE ≤ 0 days –

followed by windowing to TTE ≤ 180 days. To reduce repeated dosing artifacts, we kept the shortest TTE per patient×drug, trimmed outliers outside [Q1 – 1.5×IQR, Q3 + 1.5×IQR], and for phenotype-level summaries further kept the shortest TTE per patient. GBFDE is retained in phenotype-level displays; in per-drug facet plots we show the top 40 drugs by post-filter frequency for SJS/TEN, DRESS, and AGEP, while the “All Drugs” benchmark (diamond/line) is computed from the entire filtered set for that phenotype (not only the top-N).
