## Supplementary figures and images for "Demographics, Overlap, and Latency of Severe Cutaneous Adverse Reactions in an FDA Database"

### Supplemental Figures

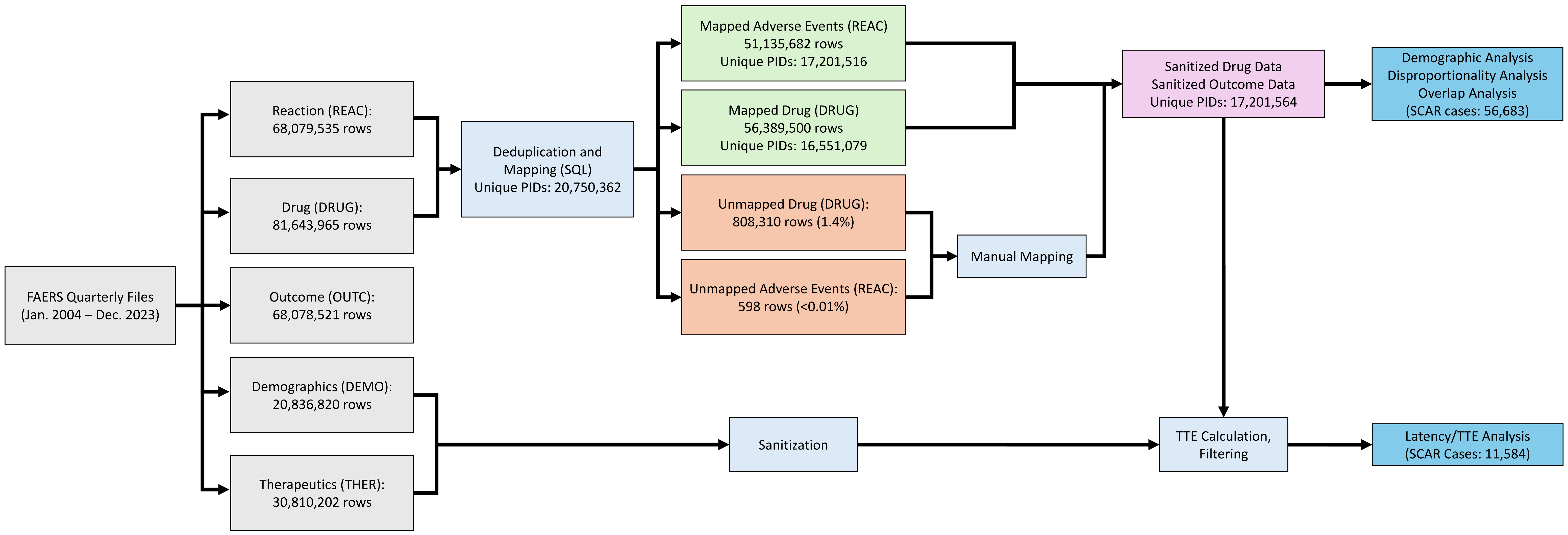

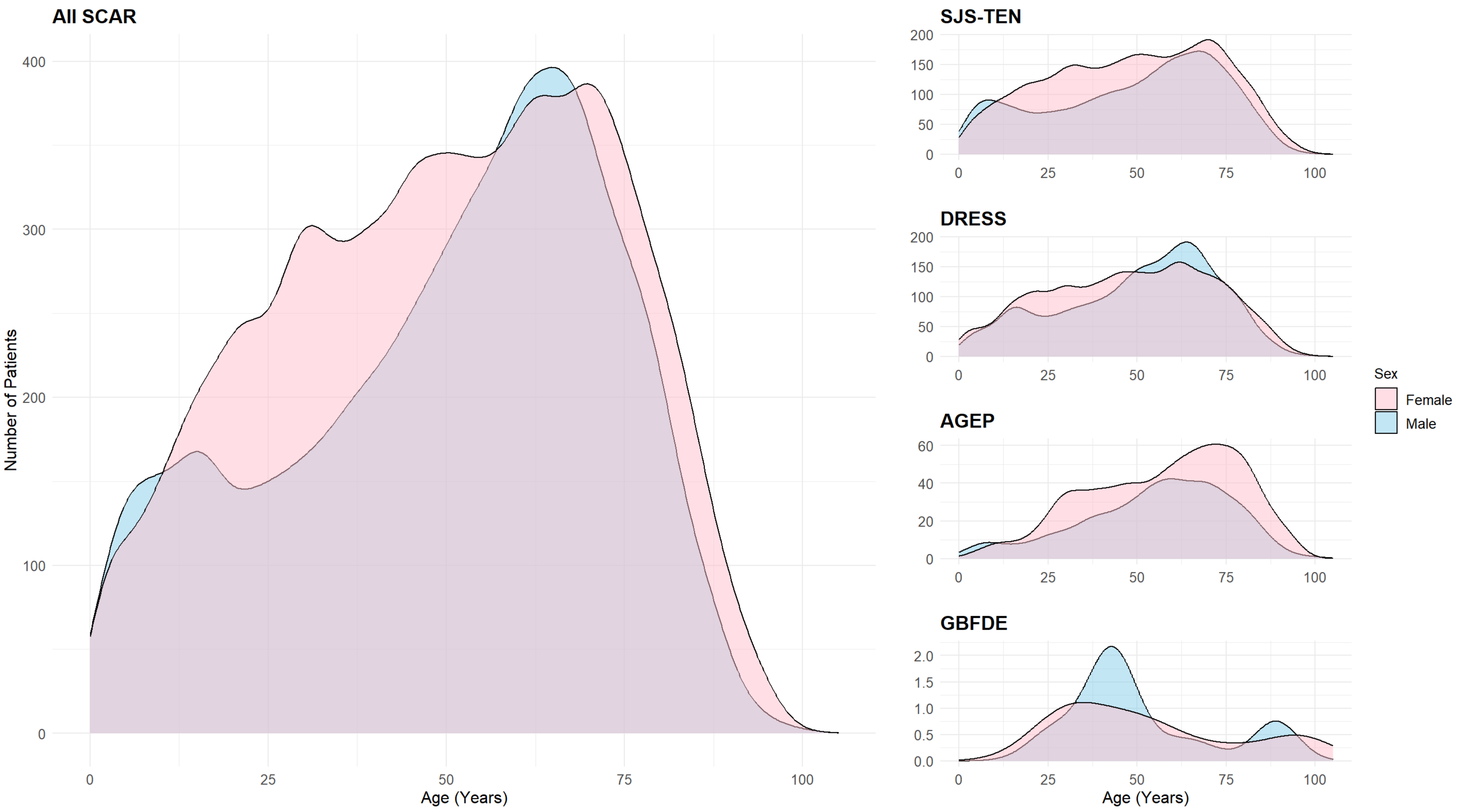

**Total SCAR**

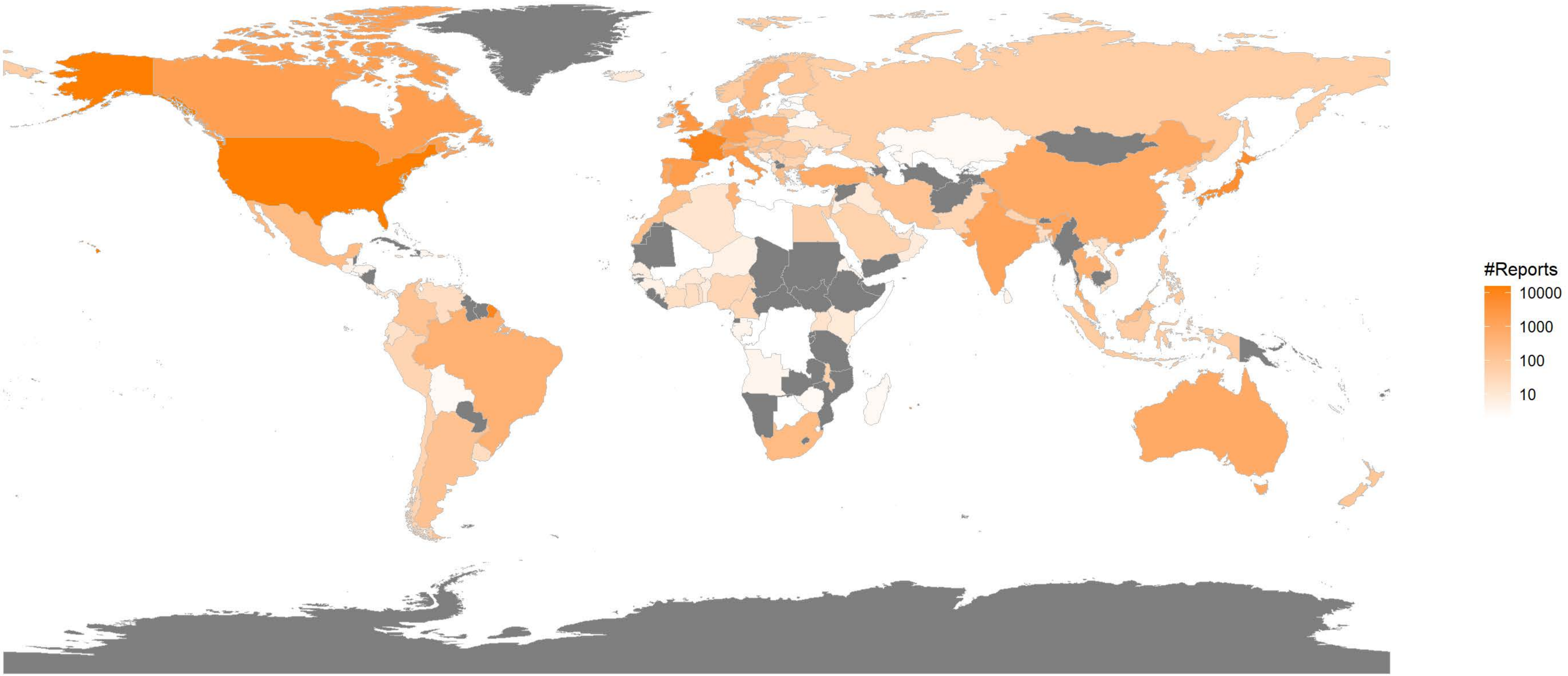

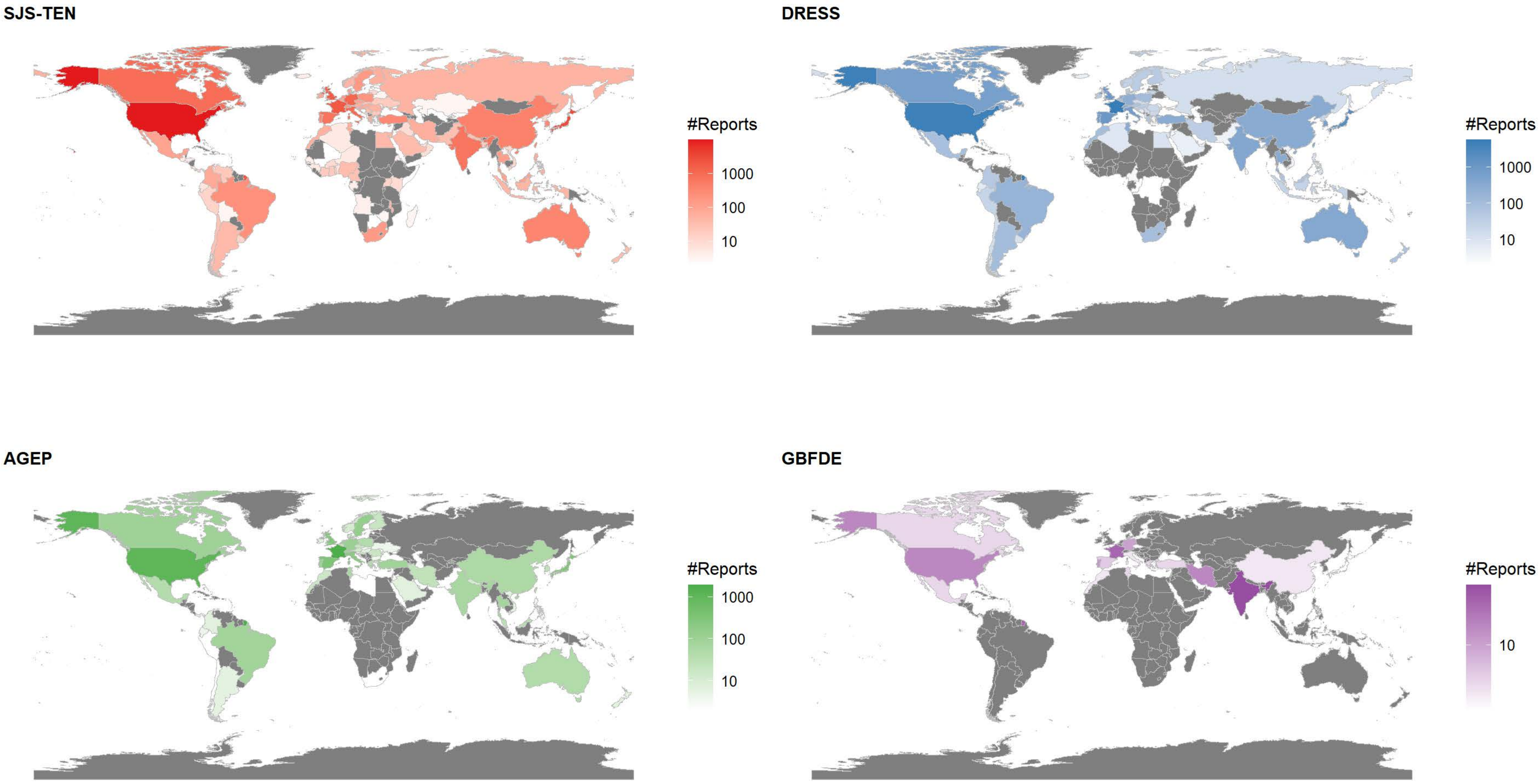

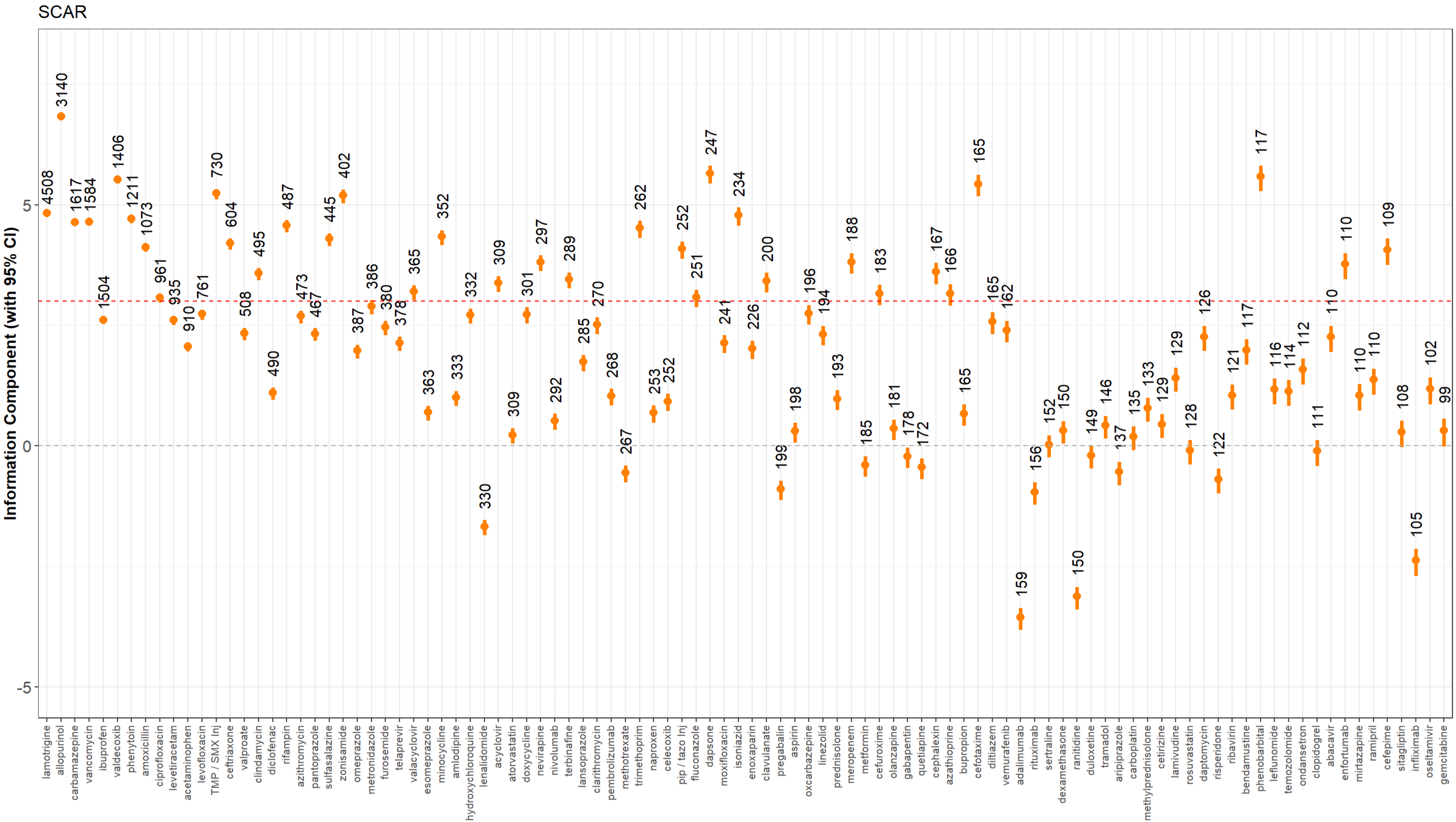

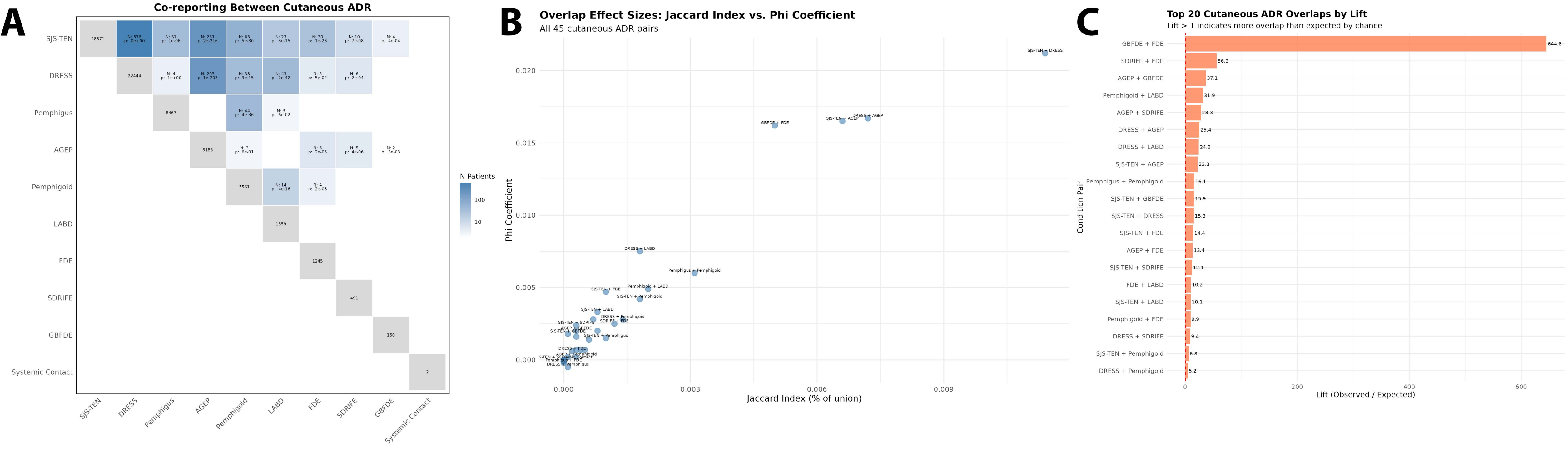

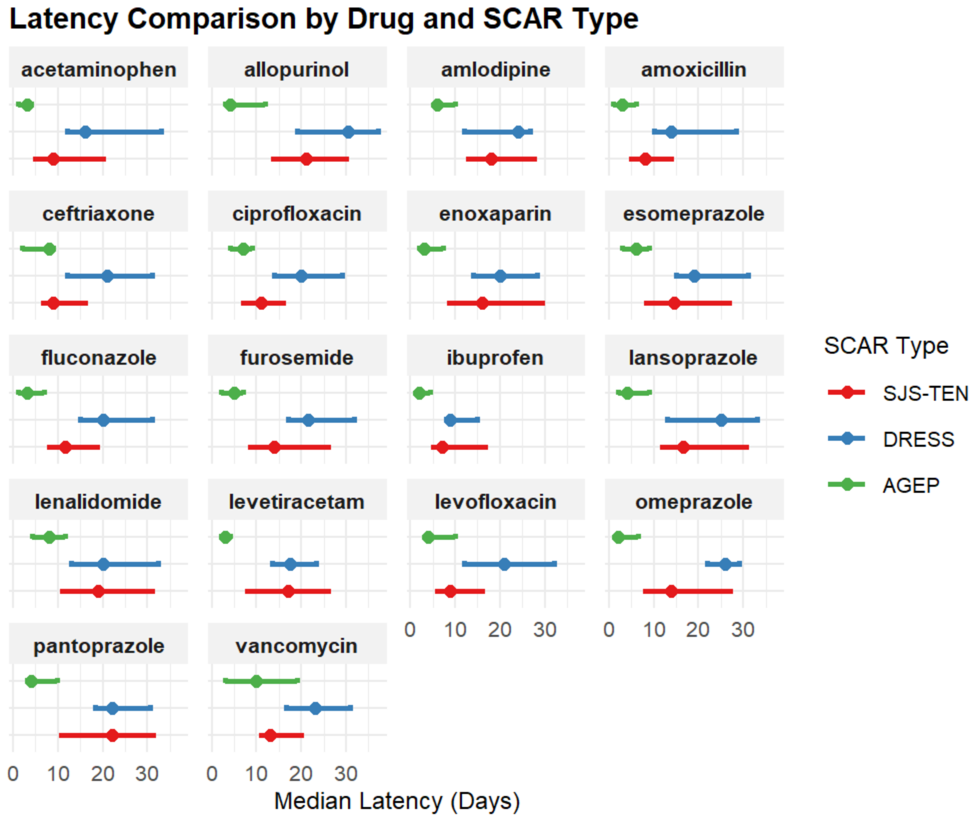
